# A Randomized, Double-Blind, Placebo-Controlled, Dose-Response Phase 2a Study of the Efficacy and Safety of a Bispecific Fusion Protein (MEDI7352) Targeting NGF and TNFα in Patients with Painful Diabetic Neuropathy

**DOI:** 10.1101/2025.11.27.25341159

**Authors:** Kirsten M. Scott, Ola Adamson, Tharani Chessell, Edward Emery, Ali Guermazi, David Howe, Richard Jenkins, Jenny Lamport, Mene Pangalos, Thomas Schnitzer, Keith Tan, Isabelle Pouliquen, Fraser Welsh, Thor Ostenfeld, Ekaterina Podchufarova, Mark A. Pilling, Nigel Brayshaw, Iain Chessell

## Abstract

Many patients with painful diabetic neuropathy (PDN) do not achieve meaningful pain relief with standard of care. MEDI7352 is a unique bispecific fusion protein targeting two key mediators of pain, TNFα and NGF (at relatively low levels of suppression). We report a randomized, double-blind, placebo-controlled, Phase-2a study (NCT03755934/EudraCT-2018-002523-42) assessing efficacy and safety of MEDI7352 in patients aged ≥18 years with inadequately controlled PDN. Patients remained on background standard-of-care treatment and were randomized to placebo (n=54) or MEDI7352 intravenously (5μg/kg [n=6], 150μg/kg [n=16], 450μg/kg [n=36]). Primary endpoint was change in pain scores from baseline to Week 12 vs placebo on a numeric rating scale (NRS). Key secondary outcomes included change in pain scores from baseline at visits prior to Week 12 and responder rates. Of 112 patients randomized, 107 received ≥1 dose study medication; mean age (standard deviation [SD]) in the modified-intent-to-treat population was 60.5 (9.34) years; 62.6% males, 37.4% females. With MEDI7352 450μg/kg, change from baseline in pain scores at Week 12 vs placebo was: −1.39 [95% CI: −2.19 to −0.58], *p*=0.0009; at Week 12, mean (SD) pain scores decreased vs baseline by −2.70 (2.08); 66.7% of patients experienced decreased pain by ≥30% (*p*=0.0311 vs. placebo); 42.4% experienced decrease ≥50% (*p*=0.0029). MEDI7352 safety was similar to placebo; no adjudicated Rapidly Progressing Osteoarthritis (RPOA) type-1 or RPOA type-2 events were observed in any group. MEDI7352 achieved statistically significant and clinically meaningful pain reduction in patients with PDN vs placebo and provides an opportunity to change the paradigm for challenging-to-treat pain.

## INTRODUCTION

An estimated 529 million people were living with diabetes in 2021, and this number is expected to more than double to about 1.31 billion by 2050 (*1*). Pain is prevalent in around 10% to 20% of patients with diabetes, and in around 40% to 50% of those with diabetic neuropathy (*2*). The majority have features of chronic sensorimotor neuropathy (*2–5*). Painful diabetic neuropathy (PDN) is often experienced as a burning, stabbing, pricking or aching sensation that is chronic and difficult to manage; it carries a substantial physical, social, and economic burden. Patients frequently experience anxiety, depression, and sleep disturbance. It is estimated that almost half of these patients do not receive any treatment for their pain.

First-line treatments for PDN include tricyclic antidepressants, pregabalin, gabapentin, and duloxetine; other treatments include opioids, topical treatments (lidocaine and capsaicin), and non-pharmacological treatments including transcutaneous electrical nerve stimulation and acupuncture. All of these treatments have limitations, and variable efficacy and responder rates; many carry serious safety concerns and the potential for abuse and dependence. With standard therapies, the number needed to treat to achieve at least 50% pain relief is around 5 to 6 (*6, 7*). Despite the poly-pharmacy approach to PDN management, fewer than 20% of patients can expect to achieve meaningful pain relief with current treatment paradigms. There is therefore a high need for new drugs for the treatment of PDN that offer an improved balance of pain relief and tolerability.

MEDI7352 is a novel bispecific fusion protein uniquely designed to engage two key biological mediators, Nerve Growth Factor (NGF) and Tumor Necrosis Factor-α (TNFα), both of which are considered to play an important role in sensitization of the nervous system and in pain pathophysiology (*8–12*). Tumor necrosis factor receptor blockers have a well-established role for disease modification in the management of rheumatoid arthritis and other inflammatory joint diseases, in which pain is often an important component (*13*); TNFα blockers are thought to be at least partially efficacious in clinical neuropathic pain (*10, 14*). For anti-NGF antibodies, several clinical studies have demonstrated efficacy in osteoarthritis (OA) and chronic lower back pain (*15–22*). However, despite significant analgesic benefits, studies have been limited to lower doses – partly due to the rare but serious occurrences of rapidly progressive osteoarthritis (RPOA), resulting in earlier-than-expected joint replacements for a small number of patients. This condition appears to be dose-related and exacerbated by co-administration of non-steroidal anti-inflammatory drugs (NSAIDs). Furthermore, RPOA is only observed after chronic dosing, appearing after approximately 24 weeks of exposure, with an unclear peak incidence (*23*). These effects have largely been responsible for previous terminations of further development of anti-NGF therapies for OA in humans. Anti-NGF therapies are approved for OA pain in dogs and cats (bedinvetmab and frunevetmab, respectively), with an adequate safety profile (*24*) and positive post-approval data showing improvements in quality of life (*25*).

The mechanism underlying the observed joint safety events remains poorly understood (*26*). One possible explanation could be NGF-related effects on cartilage formation and bone repair (*27*). There is pre-clinical evidence supporting this hypothesis: signaling via the NGF receptor promotes repair and reduces inflammation in osteoarthritic joints in mice via inhibition of nuclear factor kappa-light-chain-enhancer of activated B cells (NF-κB) (which mediates the induction of TNFα). Blocking this pathway resulted in increased NF-κB, leading to impaired bone formation and increased bone resorption (*28*). Notably, anti-TNFα therapy has shown potential to protect joints in patients with rheumatoid arthritis, limiting radiographic joint damage and the progression of structural damage (*29*).

MEDI7352 is the first bispecific, biologic molecule to be tested in chronic pain indications. Its novel mechanism, targeting both the TNFα and NGF signaling pathways, distinguishes it from regular anti-NGF approaches and may therefore offer a differentiated safety profile as well as the potential for improved efficacy. We also hypothesized that the bispecific approach would provide improved analgesic efficacy with correspondingly lower NGF suppression than molecules targeting NGF alone.

Administration of MEDI7352 produced a significant antihyperalgesic effect in pre-clinical *in vivo* models of inflammatory and neuropathic pain, at doses at which its individual separate components remained inactive. Target engagement pharmacokinetics/pharmacodynamics (PK/PD) modelling of preclinical data suggested that MEDI7352 may be able to achieve efficacy by suppressing less than 10% of circulating NGF, resulting from synergy achieved through sequestration of both NGF as well as TNFα (*30*). MEDI7352 therefore represents a potential analgesic mechanism that could avoid unwanted effects associated with higher levels of NGF suppression (hence improving safety), while maximizing analgesic benefits (*30*).

We also previously assessed MEDI7352 in a randomized, double-blind, placebo controlled, interleaved single ascending dose (SAD) and multiple ascending dose (MAD), multi-center Phase 1 study in participants with painful osteoarthritis of the knee (NCT02508155; EudraCT identifier 2015-000267-13) (*31*). While the primary purpose of that study was to investigate safety, tolerability, PK and PD of MEDI7352, exploratory endpoints also included clinical PD measures of pain. Numeric rating scale (NRS) pain scores and Western Ontario and McMaster Universities Osteoarthritis Index (WOMAC) pain subscale scores indicated that MEDI7352 had the potential to achieve dose-related analgesic effects when compared with placebo, in osteoarthritis; the safety profile was good, with no RPOA observed (*31*).

Here, we report the outcomes of a randomized, double-blind, placebo-controlled, four-stage Phase 2a exploratory, dose-response study (NCT03755934; EudraCT identifier 2018-002523-42) to assess the efficacy and safety of MEDI7352 in patients with PDN.

## RESULTS

### Patients

The study design is summarized in Fig S1. Altogether, 354 patients were screened; 112 patients were enrolled and randomized, and 107 patients received at least 1 dose of study medication (Fig. 1.)

**Fig. 1.**
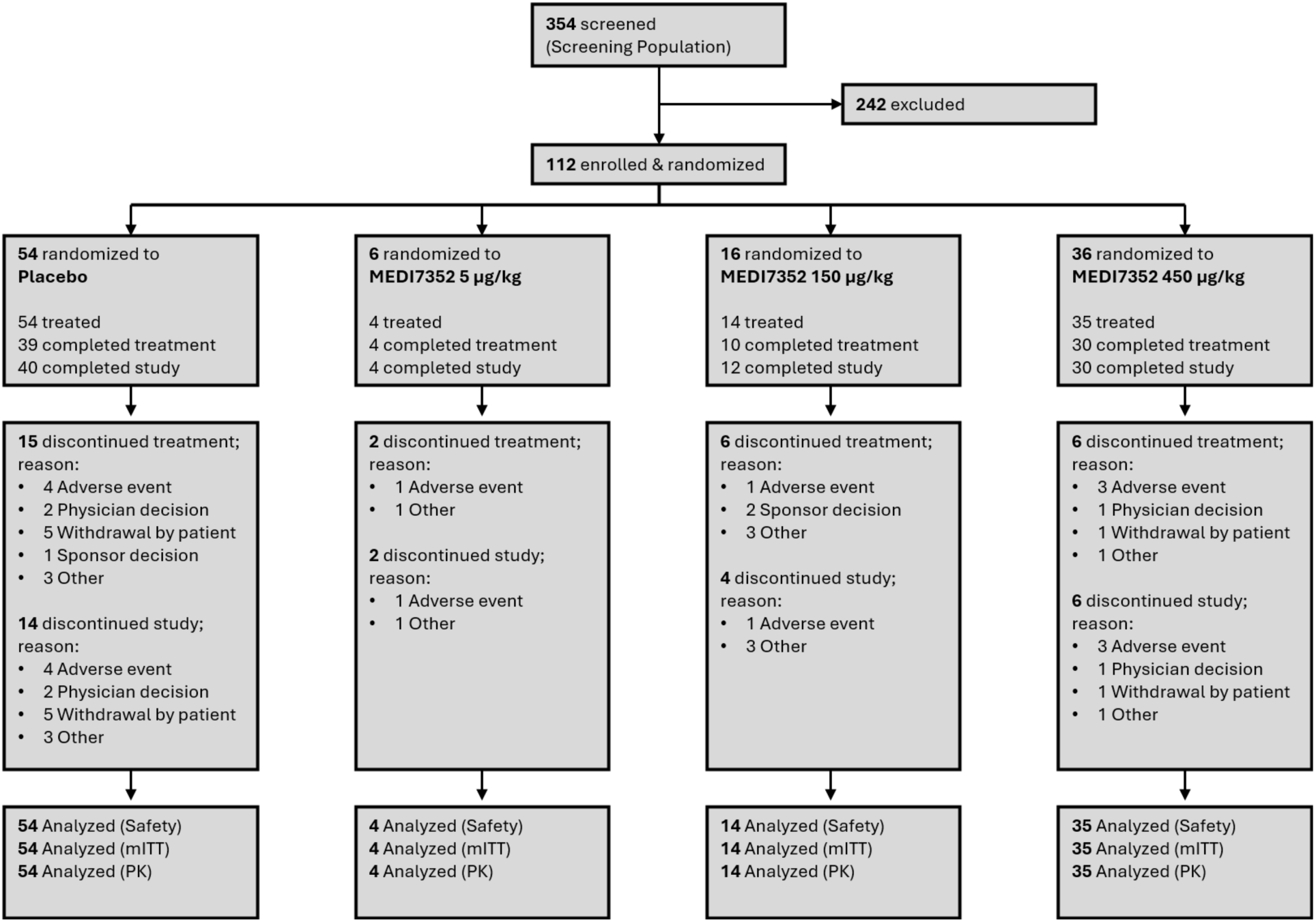
CONSORT Diagram. Screening Population includes all subjects who provided informed consent and demographic and/or baseline screening assessments. mITT = modified intent-to-treat; PK = Pharmacokinetic.

The mean (standard deviation [SD]) age of patients in the modified intent-to-treat (mITT) population was 60.5 (9.3) years with no notable differences among treatment groups (Table 1). Most patients (61.7%) were ≥18 to <65 years old. Patients were mostly male (62.6% males versus 37.4% females); 85.0% of the females were postmenopausal.

**Table 1.** Demographics and baseline characteristics (mITT/Safety Population) Prior medications were defined as medications that started before first dose of study, whether they were stopped before first dose of study medication or not. Prior medications by ATC level 2. ATC = Anatomical Therapeutic Chemical; BMI = body mass index; mITT = Modified Intent-to-treat; SD = standard deviation.

|  | Placebo<br>(N=54) | MEDI7352<br>5 µg/kg (N=4) | MEDI7352<br>150 µg/kg<br>(N=14) | MEDI7352<br>450 µg/kg<br>(N=35) | Total<br>(N=107) |
| --- | --- | --- | --- | --- | --- |
| **Demographics (mITT population)** |  |  |  |  |  |
| **Sex** |  |  |  |  |  |
| Male | 34 (63.0%) | 4 (100%) | 9 (64.3%) | 20 (57.1%) | 67 (62.6%) |
| Female | 20 (37.0%) | 0 | 5 (35.7%) | 15 (42.9%) | 40 (37.4%) |
| **Age, mean (SD), years** | 60.7 (8.02) | 62.5 (12.40) | 60.6 (11.53) | 60.0 (10.31) | 60.5 (9.34) |
| **Age category** |  |  |  |  |  |
| ≥18 to <65 years | 35 (64.8%) | 2 (50.0%) | 8 (57.1%) | 21 (60.0%) | 66 (61.7%) |
| ≥65 years | 19 (35.2%) | 2 (50.0%) | 6 (42.9%) | 14 (40.0%) | 41 (38.3%) |
| **Race** |  |  |  |  |  |
| White | 53 (98.1%) | 4 (100%) | 13 (92.9%) | 35 (100%) | 105 (98.1%) |
| Black or African American | 0 | 0 | 1 (7.1%) | 0 | 1 (0.9%) |
| Not specified | 1 (1.9%) | 0 | 0 | 0 | 1 (0.9%) |
| **Body weight, mean (SD), kg** | 91.5 (18.0) | 89.6 (13.1) | 93.3 (16.0) | 91.3 (16.9) | 91.6 (17.0) |
| **BMI, mean (SD), kg/m<sup>2</sup>** | 30.81 (5.066) | 28.28 (3.662) | 31.45 (4.774) | 31.01 (4.818) | 30.86 (4.874) |
| **Prior medications in ≥5% of Safety population** |  |  |  |  |  |
| **Analgesics** | 53 (98.1%) | 4 (100%) | 14 (100%) | 35 (100%) | 106 (99.1%) |
| Gabapentin | 17 (31.5%) | 1 (25.0%) | 11 (78.6%) | 16 (45.7%) | 45 (42.1%) |
| Pregabalin | 19 (35.2%) | 0 | 2 (14.3%) | 16 (45.7%) | 37 (34.6%) |
| Paracetamol | 7 (13.0%) | 2 (50.0%) | 3 (21.4%) | 4 (11.4%) | 16 (15.0%) |
| Duloxetine | 8 (14.8%) | 1 (25.0%) | 2 (14.3%) | 3 (8.6%) | 14 (13.1%) |
| Duloxetine hydrochloride | 10 (18.5%) | 0 | 0 | 4 (11.4%) | 14 (13.1%) |
| Amitriptyline | 7 (13.0%) | 1 (25.0%) | 2 (14.3%) | 1 (2.9%) | 11 (10.3%) |
| **Drugs used in diabetes** | 51 (94.4%) | 4 (100%) | 14 (100%) | 35 (100%) | 104 (97.2%) |
| Metformin hydrochloride | 23 (42.6%) | 0 | 4 (28.6%) | 23 (65.7%) | 50 (46.7%) |
| Metformin | 12 (22.2%) | 3 (75.0%) | 6 (42.9%) | 3 (8.6%) | 24 (22.4%) |
| Gliclazide | 5 (9.3%) | 3 (75.0%) | 4 (28.6%) | 6 (17.1%) | 18 (16.8%) |
| Insulin aspart | 10 (18.5%) | 0 | 1 (7.1%) | 7 (20.0%) | 18 (16.8%) |
| Insulin glargine | 11 (20.4%) | 0 | 2 (14.3%) | 4 (11.4%) | 17 (15.9%) |
| Empagliflozin | 6 (11.1%) | 0 | 0 | 6 (17.1%) | 12 (11.2%) |
| Insulin human | 5 (9.3%) | 0 | 3 (21.4%) | 3 (8.6%) | 11 (10.3%) |
| Insulin lispro | 6 (11.1%) | 0 | 2 (14.3%) | 3 (8.6%) | 11 (10.3%) |
| Glimepiride | 5 (9.3%) | 0 | 1 (7.1%) | 3 (8.6%) | 9 (8.4%) |
| Insulin degludec | 6 (11.1%) | 0 | 0 | 3 (8.6%) | 9 (8.4%) |
| Insulin human injection, isophane | 5 (9.3%) | 0 | 0 | 2 (5.7%) | 7 (6.5%) |
| Insulin human; insulin human injection, isophane | 5 (9.3%) | 0 | 0 | 2 (5.7%) | 7 (6.5%) |

In the mITT population, 17 patients (15.9%) had a diagnosis of OA, which mostly affected the spine (11 patients, 10.3%) and the knees (5 patients, 4.7%). Most of the patients with OA were randomly assigned to the placebo group (13 patients, 24.1%); none of the patients in the MEDI7352 150 μg/kg group had OA. Osteoarthritis was diagnosed in 1 of 4 patients (25.0%) in the MEDI7352 5 μg/kg group and 3 of 35 patients (8.6%) in the MEDI7352 450 μg/kg group. However, in line with the study exclusion criteria, the OA was not considered clinically significant in any of the patients.

Of all 107 patients in the safety population, 100 patients (93.5%) had type 2 diabetes mellitus, 6 patients (5.6%) had type 1 diabetes mellitus, and 1 patient (0.9%) had unspecified diabetes mellitus.

Analgesics and drugs used in diabetes were administered as prior medication to all patients in the MEDI7352 groups, and almost all patients in the placebo group (N=54; 53 patients received analgesics [98.1%], 51 patients received drugs used in diabetes [94.4%]) (Table 1). The most commonly reported analgesic medications were gabapentin, pregabalin, paracetamol, duloxetine, and amitriptyline, and the most commonly reported drugs used in diabetes were metformin, gliclazide, empagliflozin, and a form of insulin.

The same proportion of patients who took analgesics and drugs used in diabetes as prior medication also took them as concomitant medication during the course of the trial: 100% of patients in all MEDI7352 groups took medications in both drug classes; in the placebo group, 98.1% of patients took analgesics, and 94.4% took drugs used in diabetes (Table S1).

In terms of the disease under study (PDN) and concomitant diseases, age, body mass index, prior and concomitant medications, the recruited population adequately represented the target population for MEDI7352, and was appropriate for the type of study; demographics and baseline characteristics were either similar among the treatment groups, or the observed small differences were not considered to have a potential influence on results and their interpretation.

### Efficacy

The primary objective was evaluated by assessing the change in the weekly average of the average daily pain scores from the baseline week to Week 12 of MEDI7352 compared with placebo, as measured on an 11-point (0 to 10) NRS (referred to as ‘pain score[s]’ hereafter). At baseline, the mean (SD) pain score was similar in all treatment groups; in the overall population, the mean (SD) pain score was 6.69 (1.23) (Table 2). MEDI7352 450 μg/kg provided a statistically significant and clinically meaningful change from baseline in pain scores at Week 12 compared with placebo (MEDI7352 450 μg/kg vs placebo: −1.39 [95% Confidence Interval (CI): −2.19 to −0.58]; \*\*\**p*=0.0009; primary analysis of covariance analysis [ANCOVA]; Last Observation Carried Forward [LOCF]; mITT) (Fig. 2A.). For MEDI7352 150 μg/kg, change from baseline in pain scores at Week 12 compared with placebo was 0.72 (95% CI: −0.36 to 1.79); *p*= 0.1878. For MEDI7352 5 μg/kg, it was −1.96 (95% CI: −3.87 to −0.06); *p*=0.0437); however, this effect was not considered reliable due to the small sample size in this group (N=4).

**Fig. 2.**
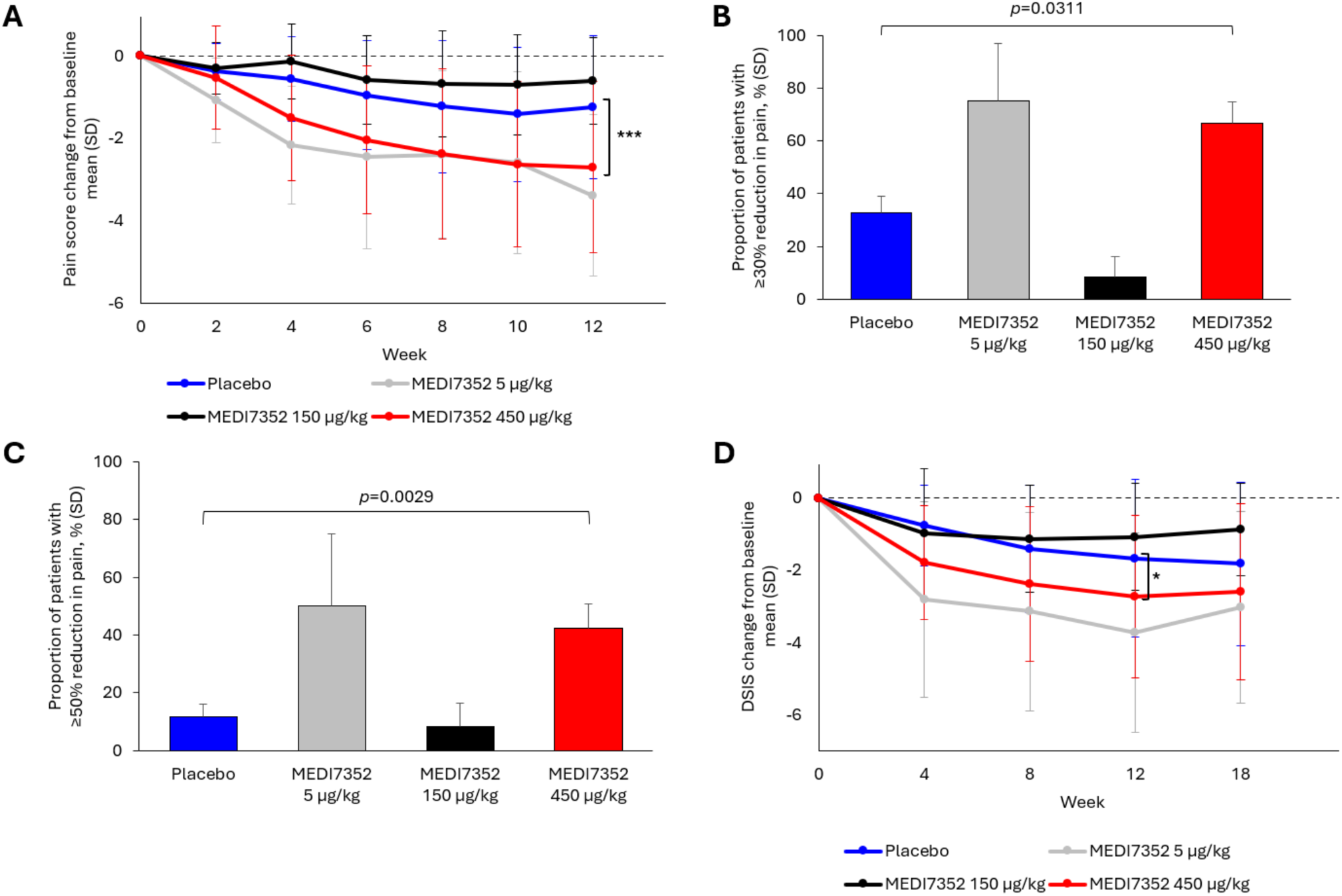
Key efficacy outcomes. A) Change in NRS pain scores from baseline (mean [SD]); MEDI7352 450 μg/kg vs placebo: −1.39 [95% CI: −2.19 to −0.58], \*\*\**p*=0.0009 (primary ANCOVA); LOCF, mITT. B and C) Percentage of patients (SD) showing ≥30% or ≥50% reduction in daily pain NRS at Week 12 compared with baseline (*p* values: Cochran-Mantel Haenszel), respectively; Observed Cases, mITT. D) Daily Sleep Interference Score; mean (SD); \**p*=0.0343 vs. placebo (mixed model for repeated measures analysis); Observed Cases, mITT. ANCOVA = analysis of covariance; DSIS = Daily Sleep Interference Score; mITT = modified intent-to-treat; NRS = Numeric Rating Scale; SD, Standard Deviation.

**Table 2.** Change in weekly average of average daily pain NRS score, compared with baseline (mITT; LOCF) [1] Baseline defined as the average of the ‘non-missing’ DPS within the seven-day period prior to randomization i.e., Day −7 to Day −1, inclusive. CFB = Change from Baseline; DPS = Daily Pain Score; LOCF = Last Observation Carried Forward; NRS = Numeric Rating Scale; SD = standard deviation.

| Change from Baseline | Placebo (N=54) | MEDI7352 5 µg/kg (N=4) | MEDI7352 150 µg/kg (N=14) | MEDI7352 450 µg/kg (N=35) | Total (N=107) |
| --- | --- | --- | --- | --- | --- |
| **Baseline [1]** |  |  |  |  |  |
| n | 54 | 4 | 14 | 35 | 107 |
| Mean (SD) | 6.61 (1.28) | 6.71 (1.81) | 6.68 (1.59) | 6.81 (0.96) | 6.69 (1.23) |
| Median | 6.93 | 7.00 | 6.57 | 6.86 | 6.86 |
| **Week 2 CFB** |  |  |  |  |  |
| n | 53 | 4 | 14 | 35 | 106 |
| Mean (SD) | -0.39 (0.69) | -1.09 (1.03) | -0.31 (0.63) | -0.54 (1.25) | -0.45 (0.92) |
| Median | -0.29 | -1.36 | -0.41 | -0.29 | -0.29 |
| **Week 4 CFB** |  |  |  |  |  |
| n | 54 | 4 | 13 | 35 | 106 |
| Mean (SD) | -0.56 (1.02) | -2.17 (1.43) | -0.14 (0.91) | -1.51 (1.51) | -0.88 (1.32) |
| Median | -0.46 | -2.21 | -0.12 | -1.14 | -0.71 |
| **Week 6 CFB** |  |  |  |  |  |
| n | 50 | 4 | 13 | 34 | 101 |
| Mean (SD) | -0.96 (1.32) | -2.46 (2.22) | -0.58 (1.07) | -2.04 (1.79) | -1.33 (1.60) |
| Median | -0.68 | -2.55 | 0.00 | -2.13 | -1.00 |
| **Week 8 CFB** |  |  |  |  |  |
| n | 46 | 4 | 12 | 33 | 95 |
| Mean (SD) | -1.24 (1.60) | -2.39 (2.02) | -0.69 (1.28) | -2.37 (2.06) | -1.61 (1.85) |
| Median | -0.86 | -2.71 | -0.71 | -2.43 | -1.43 |
| **Week 10 CFB** |  |  |  |  |  |
| n | 44 | 4 | 12 | 33 | 93 |
| Mean (SD) | -1.42 (1.63) | -2.60 (2.20) | -0.71 (1.22) | -2.63 (1.99) | -1.81 (1.86) |
| Median | -0.79 | -2.83 | -0.57 | -2.86 | -1.45 |
| **Week 12 CFB** |  |  |  |  |  |
| n | 54 | 4 | 14 | 35 | 107 |
| Mean (SD) | -1.24 (1.73) | -3.39 (1.96) | -0.62 (1.04) | -2.70 (2.08) | -1.72 (1.96) |
| Median | -0.87 | -3.38 | -0.38 | -3.14 | -1.39 |
| **Week 18 CFB** |  |  |  |  |  |
| n | 40 | 4 | 12 | 30 | 86 |
| Mean (SD) | -1.76 (1.84) | -2.61 (2.20) | -0.55 (1.23) | -2.56 (2.12) | -1.91 (1.98) |
| Median | -1.29 | -2.43 | -0.13 | -2.76 | -1.71 |

At Week 12, the mean and the median pain scores in the placebo group had decreased from baseline by −1.24 (SD: 1.73) and −0.87, respectively (Table 2). In the MEDI7352 5 μg/kg group, the mean and the median pain scores dropped by −3.39 (SD: 1.96) and −3.38 points, respectively; again, this was not considered reliable due to the small sample size. In the MEDI7352 150 μg/kg group, the mean and the median pain scores decreased slightly less than in the placebo group, by −0.62 (SD: 1.04) and −0.38 points, respectively. In the MEDI7352 450 μg/kg group, the mean and the median pain scores decreased by −2.70 (SD: 2.08) and −3.14, respectively, compared with baseline (Figure 2A and Table 2). Pain scores gradually decreased over time in all groups during Weeks 2 to 10 (and 12), including in the placebo group, with the largest reliable impact observed in the MEDI7352 450 μg/kg group (Table 2).

Among all treatment groups (disregarding MEDI7352 5 μg/kg due to small sample size), the highest proportion of patients with ≥30% or ≥50% improvement at Week 12 was seen in the MEDI7352 450 μg/kg group, in which 66.7% of patients had a decrease of pain by ≥30% (*p*=0.0311 vs. placebo; Cochran-Mantel Haenszel), and 42.4% had a decrease of pain by ≥50% (*p*=0.0029 vs. placebo;

Cochran-Mantel Haenszel) (Fig. 2B and 2C; Tables S2 and S3). This indicates a statistically significant association between the observed improvement from baseline and the treatment.

Daily sleep interference was gradually reduced (suggesting improved sleep) in all treatment groups, including placebo. The most positive impact on sleep interference that can be considered reliable was seen in the MEDI7352 450 μg/kg group; statistical analysis for this group suggested a significant association between the observed effect and treatment at Week 12 (MEDI7352 450 μg/kg vs placebo −1.00 [95% CI: −1.93 to −0.08]; *p*=0.0343; mixed model for repeated measures analysis; Observed Cases; mITT; Fig. 2D). The effect seen in the MEDI7352 5 μg/kg group was not considered reliable, due to low sample size (Fig. 2D and Table S4).

The total score for Galer neuropathic pain scale generally decreased from baseline, and the decrease was more pronounced in the MEDI7352 groups, particularly with the 450 μg/kg dose group. However, high interindividual variability was observed; for some patients, across all treatment groups (except the MEDI7352 5 μg/kg group), an increase in the total score was seen during the study (worsening pain). Statistical analysis at Week 12 (not reported) did not suggest a significant association between the treatment and observed Change from Baseline (CFB) in any of the treatment groups (Table S5).

The proportion of patients who had ‘improved’, ‘much improved’ or ‘very much improved’ on the Patient Global Impression of Change (PGIC) increased slightly over time across all treatment groups, and the statistical analysis (not reported) did not suggest an association between treatment and changes in the PGIC assessments (Table S6).

In general, the results of 36-Item Short-form Health Survey (SF-36) did not change during the study. CFB data showed a slight improvement in the physical component score in all treatment groups; slight worsening in the mental component score was seen in all treatment groups except the MEDI7352 5 μg/kg group; change in general health (categorical data) was improved across all treatment groups, but particularly in the MEDI7352 150 and 450 μg/kg groups. However, no statistically significant differences between treatment groups were observed (not reported), except for the MEDI7352 150 μg/kg group, in which a slight improvement on mental health was estimated – which was not considered reliable due to low sample size (Table S7).

Similar proportions of patients took rescue medication in the placebo group (13.0%) and in the MEDI7352 150 μg/kg group (14.3%), with a slightly lower proportion for the MEDI7352 450 μg/kg group (8.6%) (Table S8). No patients in the MEDI7352 5 μg/kg group took a rescue medication.

### Adverse events

Treatment-emergent adverse events (TEAEs) were reported with higher incidence in patients who received MEDI7352 (69.8%) compared with patients who received placebo (55.6%), with no specific trends identified in TEAEs reported to account for the imbalance (Table 3). Most of the TEAEs were considered unrelated to the investigational product (IP). Reported TEAEs were mostly mild and moderate; severe TEAEs were reported for 1 patient who received placebo (lung abscess) and 1 patient who received MEDI7352 450 μg/kg (allodynia).

**Table 3.** Overview of Adverse Events (Safety Population) Results shown are n/N; some patients experienced multiple events. AE = adverse event; IP = investigational product; SAE = serious adverse event; SOC = System organ class; PT = preferred term; TEAE = treatment-emergent adverse event; NSD = Nervous system disorder.

|  | **MEDI7352**<br>**5 µg/kg**<br>**(N=4)** | **MEDI7352**<br>**150 µg/kg**<br>**(N=14)** | **MEDI7352**<br>**450 µg/kg**<br>**(N=35)** | **All**<br>**MEDI7352**<br>**(N=53)** | **Placebo**<br>**(N=54)** | **Total**<br>**(N=107)** |
| --- | --- | --- | --- | --- | --- | --- |
| **Overview of AEs** |  |  |  |  |  |  |
| **Patients with any AE** | 4 (100%) | 10 (71.4%) | 24 (68.6%) | 38 (71.7%) | 33 (61.1%) | 71 (66.4%) |
| **Patients with any TEAE** | 4 (100%) | 10 (71.4%) | 23 (65.7%) | 37 (69.8%) | 30 (55.6%) | 67 (62.6%) |
| Mild | 3 (75.0%) | 5 (35.7%) | 10 (28.6%) | 18 (34.0%) | 15 (27.8%) | 33 (30.8%) |
| Moderate | 1 (25.0%) | 5 (35.7%) | 12 (34.3%) | 18 (34.0%) | 14 (25.9%) | 32 (29.9%) |
| Severe | 0 | 0 | 1 (2.9%) | 1 (1.9%) | 1 (1.9%) | 2 (1.9%) |
| **Patients with any TEAE leading to discontinuation of IP** | 0 | 1 (7.1%) | 3 (8.6%) | 4 (7.5%) | 5 (9.3%) | 9 (8.4%) |
| **Patients with any SAE** | 0 | 0 | 0 | 0 | 1 (1.9%) | 1 (0.9%) |
| **Patients with TEAE reported under Nervous System Disorders** |  |  |  |  |  |  |
| **Nervous system disorders** | 3 (75.0%) | 1 (7.1%) | 6 (17.1%) | 10 (18.9%) | 8 (14.8%) | 18 (16.8%) |
| Headache | 3 (75.0%) | 1 (7.1%) | 1 (2.9%) | 5 (9.4%) | 5 (9.3%) | 10 (9.3%) |
| Paraesthesia | 1 (25.0%) | 0 | 1 (2.9%) | 2 (3.8%) | 0 | 2 (1.9%) |
| Allodynia | 0 | 0 | 1 (2.9%) | 1 (1.9%) | 0 | 1 (0.9%) |
| Burning sensation | 0 | 0 | 1 (2.9%) | 1 (1.9%) | 0 | 1 (0.9%) |
| Dizziness | 0 | 0 | 1 (2.9%) | 1 (1.9%) | 2 (3.7%) | 3 (2.8%) |
| Hypoaesthesia | 0 | 0 | 1 (2.9%) | 1 (1.9%) | 1 (1.9%) | 2 (1.9%) |
| Orthostatic intolerance | 0 | 0 | 1 (2.9%) | 1 (1.9%) | 0 | 1 (0.9%) |
| Sciatica | 0 | 0 | 1 (2.9%) | 1 (1.9%) | 0 | 1 (0.9%) |
| **Patients with most frequent TEAEs by SOC and PT (in ≥3% of all MEDI7352 patients), excl. NSDs** |  |  |  |  |  |  |
| **Infections and infestations** | 1 (25.0%) | 4 (28.6%) | 7 (20.0%) | 12 (22.6%) | 21 (38.9%) | 33 (30.8%) |
| Upper respiratory tract infection | 0 | 0 | 4 (11.4%) | 4 (7.5%) | 3 (5.6%) | 7 (6.5%) |
| Urinary tract infection | 0 | 2 (14.3%) | 1 (2.9%) | 3 (5.7%) | 2 (3.7%) | 5 (4.7%) |
| Nasopharyngitis | 0 | 0 | 2 (5.7%) | 2 (3.8%) | 11 (20.4%) | 13 (12.1%) |
| **Investigations** | 0 | 3 (21.4%) | 6 (17.1%) | 9 (17.0%) | 3 (5.6%) | 12 (11.2%) |
| Alanine aminotransferase increased | 0 | 2 (14.3%) | 0 | 2 (3.8%) | 0 | 2 (1.9%) |
| Blood glucose decreased | 0 | 1 (7.1%) | 1 (2.9%) | 2 (3.8%) | 0 | 2 (1.9%) |
| **Gastrointestinal disorders** | 2 (50.0%) | 2 (14.3%) | 3 (8.6%) | 7 (13.2%) | 11 (20.4%) | 18 (16.8%) |
| Toothache | 1 (25.0%) | 0 | 1 (2.9%) | 2 (3.8%) | 0 | 2 (1.9%) |
| **Musculoskeletal and connective tissue disorders** | 2 (50.0%) | 2 (14.3%) | 2 (5.7%) | 6 (11.3%) | 9 (16.7%) | 15 (14.0%) |
| Muscle spasms | 1 (25.0%) | 0 | 2 (5.7%) | 3 (5.7%) | 1 (1.9%) | 4 (3.7%) |
| **Metabolism and nutrition disorders** | 1 (25.0%) | 2 (14.3%) | 2 (5.7%) | 5 (9.4%) | 2 (3.7%) | 7 (6.5%) |
| Hypoglycaemia | 0 | 1 (7.1%) | 1 (2.9%) | 2 (3.8%) | 1 (1.9%) | 3 (2.8%) |
| **Injury, poisoning and procedural complications** | 0 | 1 (7.1%) | 3 (8.6%) | 4 (7.5%) | 3 (5.6%) | 7 (6.5%) |
| Infusion-related reaction | 0 | 0 | 2 (5.7%) | 2 (3.8%) | 0 | 2 (1.9%) |
| **Respiratory, thoracic and mediastinal disorders** | 1 (25.0%) | 1 (7.1%) | 1 (2.9%) | 3 (5.7%) | 1 (1.9%) | 4 (3.7%) |
| Oropharyngeal pain | 0 | 1 (7.1%) | 1 (2.9%) | 2 (3.8%) | 1 (1.9%) | 3 (2.8%) |

A serious adverse event was reported for 1 patient in the placebo group, who experienced severe lung abscess (not related to IP). No TEAE was life-threatening or fatal. Nine patients (8.4%) discontinued IP due to a TEAE: 5 in the placebo group (9.3%) and 4 in the All MEDI7352 group (7.5%). Two patients in the All MEDI7352 group developed 3 adverse events of special interest of infusion-related reaction, including 1 patient in whom this event led to discontinuation of treatment with the IP. There were no joint safety adverse events of special interest, and no adjudicated RPOA type 1 or RPOA type 2 events were observed in any group.

The most commonly reported TEAEs were reported under the “Infections and infestations” system organ class (SOC) and included events such as upper respiratory tract infection, urinary tract infection and nasopharyngitis. These events were reported in a smaller proportion of patients who received MEDI7352. Common TEAEs under the “Nervous system disorders” SOC, such as headache, were reported with similar incidence between the placebo group and the All MEDI7352 group; however, paraesthesia was reported only in the All MEDI7352 group, not in the placebo group (2 patients vs 0 patients). Other common events included events under the “Gastrointestinal disorders” SOC, such as diarrhoea, which were more common in the placebo group. Events reported with a ≥3.0% greater incidence in the All MEDI7352 group than in the placebo group were muscle spasms, alanine aminotransferase increased, blood glucose decreased, infusion related reaction, paraesthesia, and toothache.

No clinically relevant observations or trends were found in the clinical chemistry, haematology, coagulation, urinalysis, vital signs, and electrocardiogram evaluations. Results of physical and neurological exams, Total Neuropathy Score-Nurse, motor and sensory nerve conduction studies and strength and tendon reflexes were in concordance with usual findings in patients with PDN. No relevant changes were observed in these parameters throughout the study.

Evaluation of hypersensitivity/anaphylactic reactions, injection-site reactions, and infusion-related reactions, drug-induced liver injury and infection risk did not raise any concerns.

Overall, the safety profile of MEDI7352 was similar to that of placebo, with no unexpected safety findings.

### Pharmacokinetic, pharmacodynamic, immunogenicity and dose-response assessments

A descriptive summary is included in the Supplementary Materials. Of note, the prevalence of anti-drug antibodies (ADAs) in this study was high, with 75.5% of patients having treatment-emergent ADAs. The presence of ADAs appeared to impact both the PK and PD of MEDI7352 (Fig. S2–S5 and Table S9). Only relatively limited PK samples were collected in this study for each patient; these data were therefore analysed by population PK methods instead – as part of a combined population PK analysis of several MEDI7352 studies (*32, 33*). There was no clear impact of ADA on downstream efficacy or safety, despite the impact on PD.

## DISCUSSION

We studied MEDI7352, a novel bispecific fusion protein targeting NGF and TNFα, and showed that at a dose of 450 μg/kg it provided a statistically significant and clinically meaningful reduction of pain in addition to standard of care, compared with placebo, 12 weeks after initiation of treatment. Pain decreased gradually over time, and for each dose this was approximately linear over time, independent of co-medication type or baseline pain level; the low numbers of patients in some of the treatment groups limited our ability to detect a dose-response. The benefit also appeared to persist 4–6 weeks after the final dose. More than 60% of patients who received the dose of 450 μg/kg reported at least a 30% decrease in pain, and more than 40% of patients receiving this dose reported at least a 50% decrease in pain. We also observed an improvement in sleep that is considered clinically meaningful (*34*). These results suggest that targeting both NGF and TNFα delivers significant efficacy (in addition to standard of care medication) that is sustained beyond the half-life of the drug (*31–33*). The mechanism underlying this remains unclear, but it implies that there is a lasting effect that is initiated by the suppression of these pathways.

This was a difficult-to treat population of patients, with significant ongoing pain who were already refractory to standard of care medications. Inadequate response to pharmacological treatments is a considerable unmet need in patients with neuropathic pain; modest efficacy, large responses to placebo, heterogeneity in diagnostic criteria, and suboptimal profiling of phenotype likely account for modest trial outcomes (*35*). Notably, in our study there was no washout period for prior pain medications before randomization, and all groups (including the placebo group) continued to receive other pain medications as part of their background standard-of-care. The improvement we observed in pain scores is consistent with observations from our previous Phase 1 study in participants with painful OA of the knee (*31*).

The safety profile of MEDI7352 was similar to that of placebo, with no unexpected safety findings. RPOA and destructive changes in the joints have been identified as a dose-related class effect for anti-NGF monoclonal antibodies (*27, 36*). No patient experienced a TEAE of OA or other joint degenerative disease during the study, and no predefined adverse events of special interest were identified in this category. Patients had been excluded if they had clinically significant OA affecting a major joint in the upper extremity, lower extremity, or axial spine, or were at risk of RPOA; diagnosis of OA was guided by American College of Rheumatology (ACR) criteria with radiologic assessment at the discretion of the investigator. However, because there was no protocol-mandated imaging in the study after screening, it is possible that there were undetected cases of RPOA1. Furthermore, given that RPOA is usually observed after 24 weeks of exposure, the duration of this study was not long enough to rule this out; we did not observe any case of subchondral insufficiency fracture in any of the large joints.

Anti-NGF monoclonal antibodies have been associated with adverse effects such as sensory abnormalities (including paraesthesia, hypoesthesia or burning sensation) in clinical studies (*15, 16, 26*), and with effects on autonomic nervous system and sympathetic dysfunction in non-clinical studies. While the incidence of such events in the present study was generally higher among MEDI7352-treated patients than in placebo-treated patients, the absolute number of patients experiencing such events was small. We hypothesise that the lower dose (and consequently, lower levels of NGF suppression) with MEDI7352 may result in fewer such adverse effects when compared with using an anti-NGF monoclonal antibody alone, at a higher dose – while still maintaining a clinically meaningful benefit.

Anti-TNFα therapies have been previously associated with an increased risk of serious or severe infections (*37, 38*). Infections and infestations were commonly reported TEAEs in this study; this included common infections such as upper respiratory tract infection, urinary tract infection, and nasopharyngitis, which were all slightly more frequent in the placebo group. Overall, we observed no increased risk of infection due to MEDI7352, but again this is more likely to be observed with more chronic dosing – and is potentially a concern in a population of patients with diabetes who are more vulnerable to infection.

As with all biologics, there may be risks of immunological reactions including hypersensitivity reactions, injection site reactions, and the consequences of immunogenicity. Fewer than 4% of patients exposed to MEDI7352 developed an infusion-related reaction, compared with 0% in the placebo group; the reported events were mild or moderate, and patients recovered on the same or on the next day. Most of the patients treated with MEDI7352 did not experience an injection site reaction.

We also compared the safety data from this present study with safety data from three other trials (not reported): a Phase 1 study to evaluate safety, tolerability, PK and immunogenicity of MEDI7352 in healthy volunteers (NCT04770428), a Phase 1 study of MEDI7352 in patients with painful OA of the knee (NCT02508155), and a Phase 2 study of the efficacy and safety of MEDI7352 in patients with painful OA of the knee (NCT04675034). This analysis showed there was general consistency with the findings in the present study. MEDI7352 was generally well tolerated across all four studies; all adverse events have been reported on clinicaltrials.gov. Of note, in the Phase 2 trial in patients with painful OA of the knee (NCT04675034), there were two events of rapidly progressive osteoarthritis type 1 (RPOA1) in the placebo group (one left knee, one right knee), one event of RPOA type 1 in the MEDI7352 Dose Level 4 group (left knee), and one of a subchondral insufficiency fracture in the MEDI7352 Dose Level 2 group (left hip). There were no overall differences in the incidence of ‘infections and infestations’. We also noted an increase in peripheral sensory symptoms (paraesthesia/hypoesthesia) in the OA study that are well described in other NGF programs and were self-limiting. There was one case of a new neuropathy that was treatment-emergent in the MEDI7352 Dose Level 4 group (NCT04675034).

In this study, we observed a high prevalence of ADAs – consistent with observations in other studies of MEDI7352. The presence of ADAs appeared to impact both PK and PD of MEDI7352, and likely confounded our ability to accurately assess free NGF suppression. A detailed population PK/PD modelling analysis of MEDI7352 PK and total NGF serum concentrations indicates that at the dose of 450 μg/kg once every two weeks for 12 weeks, predicted levels of free NGF suppression were approximately 50% – substantially lower than the predicted levels of free NGF suppression in other studies of anti-NGF antibodies in PDN (*32, 33*); a post hoc analysis indicated that we observed no correlation between ADA levels and lack of efficacy, at least with this limited dataset.

The study was disrupted by Covid-19 (resulting in a temporary pause), and a notable limitation is the relatively low number of patients enrolled, which also impacted our ability to detect a dose-response. Overall, 53 patients were exposed to MEDI7352, with only 4 patients included in the analysis sets for MEDI7352 5μg/kg. Therefore, any conclusions on safety need to be evaluated from the perspective of a relatively low sample size and short study duration of 12 weeks.

### Conclusions

MEDI7352 is a novel bispecific fusion protein designed to target two key mediators in pain signalling pathways, NGF and TNFα. This unique mechanism has previously demonstrated synergy in pre-clinical models of both inflammatory and neuropathic pain, at unexpectedly low dose; a previous Phase 1 study in patients with painful OA of the knee indicated potential for analgesic efficacy. Combining these data suggest the potential for a paradigm-changing treatment for patients with chronic pain, without the previous dose-limiting safety concerns of the anti-NGF mAb approaches.

MEDI7352 at a dose of 450 μg/kg every two weeks over a treatment period of 12 weeks provided a statistically significant and clinically meaningful reduction of pain compared with placebo, in patients who were on a background of permitted standard-of-care concomitant analgesic medications. The majority of patients in this group had at least a 30% decrease of their pain, with a significant proportion also experiencing a 50% reduction in pain. Overall, the safety profile of MEDI7352 was similar to that of placebo, with no unexpected safety findings. MEDI7352 has so far not shown a propensity to be associated with joint-related adverse events as observed with anti NGF monoclonal antibodies, but this should be caveated to note that joint-related adverse events tend to be seen after 24 weeks of dosing. The significant pain reduction provided by MEDI7352 in patients with PDN (who remained on standard of care) could potentially change the treatment paradigm for this difficult-to-treat population.

## MATERIALS AND METHODS

### Study design

This was a randomised, double-blind, placebo-controlled, exploratory, Phase 2a, multicentre study of MEDI7352 in patients aged 18 years and older with moderate to severe chronic PDN, persistent for 6 months or longer, not adequately controlled by standard of care treatments, caused by type 1 or type 2 diabetes mellitus. Trial registration: https://clinicaltrials.gov/study/NCT03755934 (date first submitted: 31 October 2018). Patients or the public were not involved in the design, conduct or reporting of this trial.

The study was conducted at 48 sites in 6 countries (the United Kingdom, Hungary, Poland, Romania, Denmark, and Spain; no patients were enrolled in Denmark or Spain), in a hospital setting. First subject enrolled November 2018, last subject last visit June 2023.

The study was performed in accordance with the ethical principles aligned with the Declaration of Helsinki, and in compliance with International Council for Harmonisation (ICH) Good Clinical Practice, including the archiving of essential documents. Written informed consent was obtained from all patients prior to enrolment.

The primary objective was to assess the efficacy of MEDI7352 vs placebo on chronic pain in patients with PDN currently taking standard of care medication for their PDN pain. The secondary objectives were to assess safety, tolerability, pharmacokinetics and pharmacodynamics, immunogenicity, and dose-response of MEDI7352 in patients with PDN.

The study incorporated a screening period of up to 45 days and a 12-week double-blind treatment period during which MEDI7352 or placebo was administered intravenously on 6 occasions, with each dose separated by 14 days. There was a 6-week follow-up period (Fig. S1).

### Patient population

Key inclusion criteria were: male, or post-menopausal or surgically sterile females; 18 to 80 years of age (inclusive) on day of randomization; body mass index of ≤42 kg/m^2^; chronic pain associated with PDN with mean pain intensity score of ≥4, as measured on an 11-point (0 to 10) NRS; willing and able to discontinue all nonsteroidal anti-inflammatory drug or cyclooxygenase2 analgesic therapy; taking medication for the treatment of PDN. Patients had to be taking at least 1 of the first-line medications (consistent with regional or local standard of care guidelines for PDN) belonging to either the anticonvulsant class (pregabalin or gabapentin) or the antidepressant class (duloxetine, venlafaxine, or amitriptyline), but no more than 1 medication from a single class. During the study, NSAIDs, analgesic doses of aspirin, and Cox-2 inhibitors were prohibited.

Key exclusion criteria were: other clinically significant neuropathy; diagnosis of clinically significant OA (affecting a major joint in the upper or lower extremity or axial spine; or other degenerative disease affecting any joint in subjects for whom, in the opinion of the investigator, there was an identified risk of osteonecrosis, RPOA, subchondral insufficiency fractures, neurogenic arthropathy, or analgesia-induced arthropathy); other chronic pain conditions; major psychiatric disorders; significant cardiovascular or lung disease; clinically important infections; other serious illnesses.

### Interventions, randomization and blinding

Four study Stages were planned (Fig. S1.). Patients in Stage 1 were to be randomly assigned to placebo or the lowest dose of MEDI7352 (5 μg/kg) until at least 10 patients had been recruited. In Stage 2, up to a maximum of 30 patients were to be randomly assigned to placebo or MEDI7352 150 μg/kg. In Stage 3, approximately 67 patients were to be randomly assigned in a 1:1 ratio to placebo or MEDI7352 450 μg/kg to ensure sufficient patients were evaluable for the pre-planned interim analysis.

Administrative analyses were conducted during Stage 3 to confirm decision-making for Stage 4 with respect to exact sample size and dose allocation ratio. Upon completing enrolment for Stage 3, an interim analysis was to be performed before Stage 4. Approximately 165 eligible patients were planned to be randomly assigned to treatment with equal allocation across 3 dose levels of MEDI7352 or placebo in Stage 4, to ensure that approximately 236 patients were evaluable for the efficacy analysis of Stages 2 to 4 combined.

After all patients in Stage 3 completed the study, the sponsor decided to prematurely terminate the study because enrolment was taking longer than anticipated, making it challenging to progress with the current protocol to Stage 4. The sponsor halted screening activities on 12 January 2023 and informed sites about the study termination on 11 April 2023. Ongoing patients continued on treatment until end of the study and the required follow-up period as per protocol. A 50 μg/kg dose planned for Stage 4 was not evaluated.

The randomization schedule was computer-generated by Premier Research before the study, using a permuted block algorithm. No one involved in the performance of the study had access to the randomisation schedule before official unblinding of treatment assignment. All patients, investigators, and study personnel involved in the conduct of the study, including data management, were blinded to treatment assignment – with the exception of a specified unblinded statistician and programmer from Premier Research, unblinded site monitors and clinical managers from Premier Research, and an unblinded pharmacist at each site. Unblinded personnel did not participate in study procedures or data analysis prior to unblinding. Data were prepared for review and interim analysis by unblinded personnel who were not otherwise involved in the conduct of the study.

### Efficacy assessments

The primary efficacy endpoint was change in the weekly average of the average daily pain scores from the baseline week to Week 12 of MEDI7352 compared with placebo, as measured on an 11-point (0 to 10) NRS. Key secondary endpoints were 1) Change in the weekly average of the average daily pain score, as measured on an 11-point (0 to 10) NRS, from baseline to Weeks 2, 4, 6, 8, and 10 of treatment and the week before the follow-up visit; 2) Percentage of patients who achieved ≥30% and ≥50% reductions in the weekly average of the average daily pain score from baseline during Weeks 4, 8, and 12 of treatment and the week before follow-up; 3) Change in Daily Sleep Interference Scale from baseline to Days 28, 56, and 84 of treatment and the follow-up visit; 4) Change in Galer Neuropathic Pain Scale from baseline to Days 28, 56, and 84 of treatment and the follow-up visit; 5) Proportion of patients who have ‘improved’, ‘much improved’, or ‘very much improved’ relative to baseline on the PGIC on Days 28, 56, and 84 of treatment and the follow-up visit; 6) Change in 36-Item Short-form Health Survey from baseline to Day 84 of treatment; 7) Change in the amount of rescue medication used (in terms of dosage/day) from baseline to Week 12 of treatment.

### Safety and adverse event assessments

Safety and tolerability assessments included: Adverse Events (AEs) and serious AEs, physical and neurological examinations, neuropathy assessments (Total Neuropathy Score-Nurse), strength (dorsiflexion) and deep tendon reflex (knee and ankle) assessments, vital signs, 12-lead electrocardiograms, clinical laboratory testing (hematology, clinical chemistry, coagulation, and urinalysis), motor and sensory nerve conduction, concomitant medication assessment, injection-site reaction assessment, and infusion reaction assessments.

### Pharmacokinetic, pharmacodynamic, immunogenicity and dose-response assessments

Pharmacokinetic parameters to be assessed for MEDI7352 were to include Cmax, time of Cmax, area under the serum concentration-time curve from time 0 to infinity, area under the serum concentration-time curve from time 0 to the time of the last quantifiable serum concentration, area under the serum concentration-time curve for the dosing interval at steady state, Cmax at steady state, volume of distribution at steady state, half-life, and total body clearance at steady state.

Pharmacodynamics assessments included NGF measurements. Other assessments included immunogenicity (ADA) and dose response.

### Statistical Analysis

Approximately 272 eligible patients were planned to be randomly assigned to double-blind treatment with 1 of 4 dose levels of MEDI7352 (5, 50, 150, and 450 μg/kg, dependent upon the stage of the study) or placebo, to ensure that approximately 236 patients were eligible for the efficacy analysis of Stages 2, 3, and 4 combined. There was no formal sample size calculation for stage 1; 10 patients in Stage 1 were considered sufficient for the initial assessment of safety. The sample size for Stages 2 to 4 combined was determined by a formal power calculation (see below) and the size of Stage 3 was defined to confirm decision-making for the Stage 4 sample size and dose allocation ratio.

This study was powered at greater than 80% to detect a statistically significant (1-sided alpha = 0.025) dose-response relationship when the true Week 12 placebo-corrected change from baseline difference at the 450 μg/kg dose was 1.25 on the 11-point NRS scale (MEDI7352-placebo treatment) and the true dose-response followed a hyperbolic maximum effect of the drug (E_max_) relationship, with the dose to achieve 50% of the maximum effect (ED50) within the range of 1 to 750 μg/kg. This calculation also assumed: 1) The true standard deviation was 2.4, based on other studies with pregabalin in PDN. 2) Data from Stages 2, 3, and 4 would be combined so that the number of patients evaluable for the dose-response analysis was 236. The total number of patients evaluable for efficacy for the placebo, 50, 150, and 450 μg/kg doses of MEDI7352 would be equal to 81, 37, 51 and 67, respectively. 3) The dose-response hypothesis test was multiplicity adjusted in order to control the type 1-error.

The study included 4 analysis populations: 1) Screening: all subjects who provided informed consent and/or assent and provided demographic and/or baseline screening assessments, regardless of their randomization and treatment status in the study. 2) Safety: all patients who received at least 1 dose of double-blind study medication. 3) mITT: used for all efficacy analyses and included all randomized patients who received at least 1 dose of double-blind study medication and had at least 1 post-baseline NRS assessment. 4) PK population included all patients for whom a PK sample was obtained and analysed. Patients receiving placebo were not included in the summary and analysis of PK parameters.

Efficacy, safety and tolerability data were summarised descriptively. Primary and secondary endpoint efficacy data were tabulated according to the ‘observed cases’ approach. In addition, if there were missing data at a key analysis time point (Weeks 4, 8, and 12), then results were also tabulated according to LOCF and baseline observation carried forward (BOCF).

The main statistical analysis of the primary efficacy endpoint at Week 12 used the multiple comparison procedure modelling approach (MCP-MOD) on LOCF data, which is a well-established statistical methodology for establishing both the existence of a dose response and modelling the underlying dose-response relationship. The underlying model was an ANCOVA with dependent variable ‘CFB to Week 12 (LOCF)’, and independent variables included: dose group as a factor variable with the placebo group as the reference level, baseline score (i.e. baseline weekly average pain [NRS]) as a continuous variable, and co-medication type.

In addition, changes from baseline in continuous endpoints were compared between treatment groups using mixed models for repeated measures including terms for treatment (as a factor), time point (as a factor), the interaction between treatment and time point, and the baseline value of the variable undergoing analysis. Binary outcomes were analysed using generalised estimating equations, with the models including the same terms as the mixed models repeated measures. Other non-binary categorical endpoints were analysed using Cochran-Mantel-Haenszel statistics for stratified data. The detailed statistical analysis plan and protocol of this trial are available at: https://www.astrazenecaclinicaltrials.com/study/D5680C00002/.

## Data Availability

Data which is not restricted by confidentiality considerations are available on reasonable request to the authors

## Acknowledgments

The authors would like to thank the participants, the site investigators, and the site staff who participated in the trials. The authors also thank Stanislav Ignatenko for Phase 1 (SAD/MAD), Jon Hatcher and Darren Schofield for their work to construct and pre-clinically characterize MEDI7352, and Andreas Leidenroth of Larkfields Ltd for medical writing support.

## Funding

The studies reported in this manuscript were funded by AstraZeneca.

## Author contributions

Conceptualization: IC, FW, TC, TO, NB, MP

Methodology: IC, FW, TC, TO, TS, AG, KT, IP, NB, EP

Investigation: FW, TC, KMS, DH, TO, TS, AG, OA, KT, IP, DH, JL, NB, EP

Funding acquisition: FW, IC, MP

Project administration: FW

Supervision: FW, IC, MP, TO, EP

Writing – original draft: FW, IC, KMS, EE

Writing – review & editing: All authors

## Competing interests

KMS, OA, TC, TO, EE, EP, DH, RJ, MP, KT, IP, FW, MAP and IC are employees or former employees of AstraZeneca and may own shares. AG is a shareholder of BICL, LLC and consultant to Novartis, TissueGene, Coval, Paradigm, Peptinov, Scarcell, Levicept, Pacira, ICM, Medipost, 4Moving and Formation Bio.

## Data and materials availability

The trial clinical study report synopsis, protocol, statistical analysis plan, and results are all available at: https://www.astrazenecaclinicaltrials.com/study/D5680C00002/

## Notes

### Competing Interest Statement

The authors have declared no competing interest.

### Clinical Trial

NCT03755934/EudraCT-2018-002523-42

### Funding Statement

This study was funded by AstraZeneca

### Author Declarations

Ethical approval was granted by all sites involved in the conduct of this study. Specifically: Bioethics Committee at the Kujawsko Pomorskie Regional Medical Chamber in Torun ul.Ignacego Danielewskiego 6, 87 to 100 Toru, Poland National Bioethics Committee for Medicines and Medical Devices Spitalul Colentina, Exterior Building C, 6, Dr.Grozovici Str., District 2, Bucharest, 021105, Romania South Central Oxford A Research Ethics Committee Bristol Research Ethics Committee Centre Whitefriars, Level 3 Block B, Lewins Mead, Bristol, BS1 2NT, United Kingdom The Health Research Ethics Committees Borgervaenget 3, stuen 2100 Kobenhavn 0, Denmark Belugyminiszterium, 1903 Budapest, Pf. 314, Hungary

